# External validation of a deep-learning mandibular ORN prediction model trained on 3D radiation distribution maps

**DOI:** 10.1101/2023.12.04.23299221

**Authors:** Laia Humbert-Vidan, Christian R Hansen, Vinod Patel, Jørgen Johansen, Andrew P King, Teresa Guerrero Urbano

## Abstract

**Background and purpose:** Mandibular osteoradionecrosis (ORN) is a severe side effect affecting patients undergoing radiation therapy for head and neck cancer. Variations in the bone’s vascularization and composition across the mandible may influence the susceptibility to ORN. Recently, deep learning-based models have been introduced for predicting mandibular ORN using radiation dose distribution maps to incorporate spatial information. These studies, however, only feature internal validation on a holdout subset of the data used for training.

**Materials and methods:** This study externally validated a 3D DenseNet-40 (DN40) ORN prediction model on an independent dataset. Model performance was evaluated in terms of discrimination and calibration, with Platt scaling applied for improved external calibration. The DN40 model’s discriminative ability on the external dataset was compared to a Random Forest model on corresponding dose-volume histogram (DVH) data.

**Results:** The overall model performance was worse at external validation than at internal validation, with Platt scaling improving balance between recall and specificity but not significantly improving the overall calibration. Although the discrimination ability of the DN40 model was slightly lower at external validation (AUROC 0.63 vs. 0.69), this was statistically comparable to that of a DVH-based RF model for the same dataset (p-value 0.667).

**Conclusions:** Our results suggest that, in addition to potential model overfitting issues, dosimetric data distribution differences between the two datasets could explain the low generalisability of the DN40 ORN prediction model. Future work will involve a larger and more diverse cohort.

## 1 Introduction

Mandibular osteoradionecrosis (ORN) is a severe late side effect that affects 4-8% [1] of patients who have undergone radiation therapy (RT) for head and neck cancer. Radiation-induced fibrosis of the irradiated tissues extends to the blood vessel walls, eventually resulting in a reduced blood supply and subsequent necrosis of the lower jawbone [2]. Depending on the severity, the clinical management of ORN may range from more conservative treatments to complex and costly surgical interventions such as segmental resection of the mandible, which are highly detrimental to the patient’s quality of life [3].

In addition to radiation dose, other risk factors have been identified, including dental extractions, pre-RT surgery to the mandible, smoking, poor oral hygiene and a sub-optimal dentition, and multiple studies [4, 5, 6, 7, 8, 9, 10] have analysed associations between these factors and the development of mandibular ORN. An increased incidence of ORN has also been observed in the HPV-associated OPC group of patients, that are typically younger, with better dental status and without the lifestyle factors associated with ORN (e.g. smoking and alcohol) [11]. Van Dijk et al. [12] developed the first ORN normal tissue complication probability (NTCP) model based on the D30% of the mandible bone and pre-RT dental extractions as predictors. Dose-volume parameters have been used clinically and in NTCP models for decades, but they have limitations [13, 14]. With such parameters, the volumetric dose distribution within an organ is reduced to a unidimensional number that does not capture potential clinically relevant spatial information. Thus, the dose-volume histogram (DVH) effectively, albeit incorrectly, assumes a heterogeneous dose distribution, organ function, and radiosensitivity within a structure. As a result, a DVH-based NTCP model might not correctly reflect the true relationship between radiation dose and toxicity for each organ sub-unit. For instance, the density and composition of the bone varies across the mandible, with some areas consisting of compact or cortical bone while others consist of less dense or trabecular bone. The lower jaw is considered more prone to ORN as the bone is denser and has reduced vascularity compared to the upper jaw. Moreover, ORN is also more likely to occur in the posterior jaw as radiation doses tend to be higher posteriorly due to its proximity to the primary tumour [15].

The use of radiation dose distribution maps as the dose information for predicting radiation-induced toxicities has been explored with deep learning (DL) methods [16, 17]. More recently, this approach has also been introduced in the prediction of mandibular ORN and compared to DVH-based approaches [18, 19, 20]. However, these studies featured only internal validation on a holdout subset of the data used for training. As described by the TRIPOD statement [21], while internal validation may inform on the model performance during its development, the external validation of a model will provide insight into how well a model may be applicable to independent datasets. Model validation studies in the context of spatial dose NTCP modelling are limited, probably due to the technical complexities involved in the data preparation process. However, external validation of such models is necessary for their acceptance in a clinical context largely dominated by DVH-based models.

The current study aimed to externally validate an existing DL-based ORN prediction model [18]. The model was developed in a UK population treated at Guy’s and St Thomas’ Hospitals (GSTT) and was externally validated in an independent dataset from a Danish population treated at Odense University Hospital (OUH) [9].

## 2 Materials and methods

### 2.1 Patient selection

The training and the external validation cohorts are described in Table 1. In both cohorts, the subjects were retrospectively selected from clinical databases using a control-case matching approach based on the primary tumour site and treatment year. The primary tumour sites include the oropharynx, oral cavity, larynx and others (paranasal sinus, salivary glands and unknown primary). The inclusion and exclusion criteria from the PREDMORN study protocol [22] were followed. Mandibular ORN severity was staged according to the Notani system [23]. However, since the prediction model was developed as a binary classification model, the ORN stage was dichotomised, and any grade of ORN was considered an ORN case.

**Table 1:** Clinical data distribution for the GSTT and OUH cohorts.

|  | GSTT | OUH |
| --- | --- | --- |
| Dataset size | 92 ORN + 92 controls | 45 ORN + 45 controls |
| Age (ORN/control) | 61 (28-83) / 60 (31-88) | 59 (38-77) / 60 (31-76) |
| Gender - male (ORN/control) | 72% / 78% | 73% / 67% |
| Primary tumour sites (matched cohort) | Oral cavity (OC) (30%)<br>Oropharynx (OP) (57%)<br>Larynx (3%)<br>Other<sup>1</sup> (10%) | OC (40%)<br>OP (56%)<br>Larynx (2%) |
| FU time (control) | 3.9 (0.4-8.4) | 3.0 (0.2-9.6) |
| Smoking (ORN/control) | 79% / 70% | 89% / 81% |
| Alcohol (ORN/control) | 77% / 78% | Not available |
| Extractions (ORN/control) | 60% / 61% | 80% / 70% |
| PORT (ORN/control) | 38% / 40% | 38% / 34% |
| Chemotherapy (ORN/control) | 64% / 60% | 29% / 28% |

### 2.2 Treatment

All subjects in both cohorts were treated with curative intent using intensity-modulated radiation therapy (IMRT). The subjects in the GSTT cohort were planned with the Monaco (Elekta AB, Stockholm, Sweden) and Eclipse (Varian Medical Systems, Milpitas, CA) treatment planning systems (TPS). Radical primary IMRT cases were prescribed a total dose of 65-70 Gy in 30-35 fractions and 55 Gy in 20 fractions in selected cases. Radical post-operative IMRT (PORT) cases were prescribed 60-66 Gy in 30-33 fractions and 50 Gy in 20 fractions in selected cases. The subjects in the OUH cohort were planned on the Pinnacle TPS and treated on the Elekta accelerators. The dose prescription was 66 or 68 Gy in 2 Gy fractions 5-6 fractions per week following the Danish Head and Neck Cancer Group (DAHANCA) national guidelines from 2013 [24, 25]. Patients were treated with simultaneous integrated boost, i.e., 60 Gy to CTV2 and 50 Gy to CTV3 (elective regions). Concomitant weekly cisplatin and radiosensitiser nimorazole were prescribed according to the DAHANCA guidelines.

### 2.3 Dosimetric data

The mandibles were segmented on the radiotherapy planning computed tomography (CT) images by each centre on their own cohort and using their respective TPS. The entire mandible structure was contoured, excluding the teeth. The reconstructed bone was not included in cases where mandibulectomy and subsequent mandible reconstruction were performed. For the training dataset, the RT dose, CT image and RT structure DICOM files, as well as dose-volume histogram data – DVH, were exported from the TPS. The external validation dataset was extracted from the Danish RT doseplan database, the DICOM Collaboration system (DcmCollab, https://dcmcollab.rsyd.dk/). The mandible 3D dose distribution maps were created by masking the clinically planned radiation dose distribution volume with the manually contoured mandible structure. Correction to an equivalent dose in 2 Gy fractions (EQD2) was applied and the dose maps were then min-max normalised to within 0 and 1 intensity values.

### 2.4 ORN prediction models

#### 2.4.1 3D DenseNet-40 Convolutional Neural Network

A 3D (DN40) [26] convolutional neural network (CNN) was trained on 3D radiation dose distribution maps of the mandible for the binary classification of ORN vs. no ORN subjects using the GSTT cohort [18]. The 3D DN40 CNN (Figure 1) consists of three dense blocks and two transition blocks, where all the convolutional, pooling, batch normalisation and dropout operations are three-dimensional. Each dense block is formed of 12 dense layers. The output from the last dense block is reduced to one dimension with a 3D average pooling layer and flattened. Finally, a fully connected layer followed by a softmax layer provides the binary classification probabilities. Implementation of the DN40 was done within the Medical Open Network for Artificial Intelligence (MONAI) (https://monai.io/) Pytorch-based framework.

**Figure 1:**
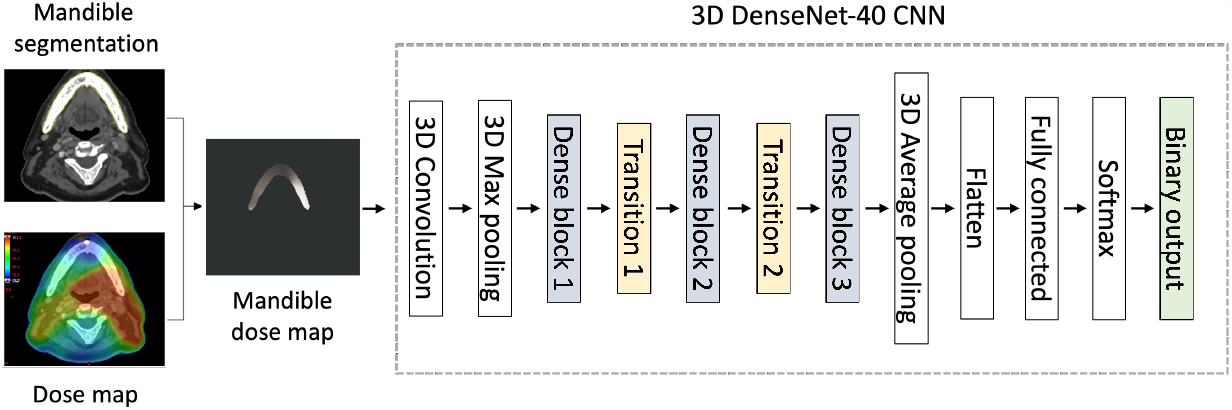
Image processing and 3D DenseNet-40 CNN (DN40) pipelines. The clinical radiation dose map was masked with the mandible segmentation to obtain the mandible dose map, which was used as the input for the DN40.

#### 2.4.2 Random Forest classifier

A random forest (RF) classifier [27] was trained on the DVH data from the external dataset and the performance compared to the DN40 model trained on the corresponding dose maps for the same dataset. The RF was implemented using the `sklearn.ensemble.RandomForestClassifier` and `model_Selection.GridSearchCV` Python modules. For the grid search CV procedure, the following hyperparameters were considered: bootstrap (True, False), maximum depth (1, 2, 10, 20, None), maximum features (auto, sqrt), minimum samples per leaf (1, 2, 4), minimum samples per split (2, 5, 10) and number of estimators (200, 500, 1000, 1300, 1700, 2000).

### 2.5 Statistical analysis

The DN40 model was internally and externally validated. In both validation processes, the model’s performance was assessed in terms of its discriminative ability and calibration.

#### 2.5.1 Internal and external validation

Internal validation was performed using a stratified nested 5-fold cross-validation (CV) approach, which consists of an inner CV loop encompassed by an outer CV loop (Figure 2). For each of the outer CV folds, hyperparameter optimisation is performed j times in an inner j-fold CV approach where the outer CV train dataset is further split into train and validation sets. Finally, for each of the outer CV folds, the entire training set is used for training using the optimised hyperparameters obtained from the inner CV, and the prediction accuracy can be calculated on the held-out test set. In this way, the test set of each outer CV fold remains completely unseen, avoiding the bias introduced in traditional CV. A k-fold CV approach is stratified when the class balance is maintained in all CV folds. The external validation process indicates how well the trained DL model generalises on an independent dataset, i.e. whether the model can produce predicted probabilities that are accurate and well-calibrated not just on the training data but also on unseen data. External validation was performed by testing the discrimination and calibration of the model on an independent dataset (OUH).

**Figure 2:**
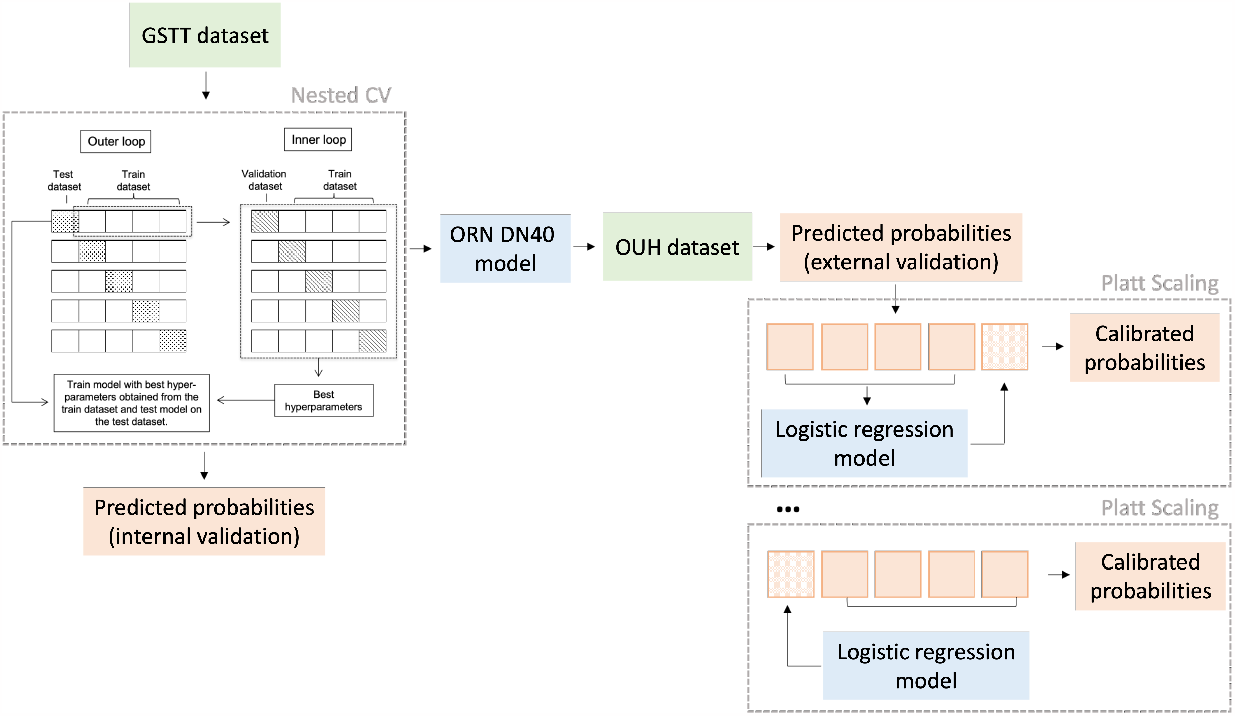
Workflow implemented to obtain the calibrated probabilities applying Platt Scaling on the raw predicted probabilities that resulted from the external validation of the DN40 ORN model on the OUH dataset. A 5-fold CV approach was followed, where Platt Scaling was applied to the test subset at each fold.

#### 2.5.2 Model discrimination

Discrimination was measured using the accuracy, recall, specificity, precision and area under the receiver operating characteristic curve (AUROC) metrics. The ROC curve was obtained by plotting the predicted probabilities for the positive class (ORN) with a probability classification threshold of 0.5 (i.e. probabilities below and above 0.5 corresponded to no ORN and ORN class predictions, respectively).

#### 2.5.3 Model calibration

Model calibration in DL classification models is crucial to ensuring that the predicted probabilities reflect the true likelihood of the predicted outcomes. Calibration-in-the-large (or mean calibration) compares the average predicted class probability to the overall event rate [28]. Alongside mean calibration, reliability diagrams were used for a visual assessment of the general over- or under-estimation of the predicted probabilities. The intercept and slope of the calibration curve provided additional information on the calibration performance. Calibration was also measured with the logarithmic loss (LogLoss). The Brier score (BS) was used to assess the overall model performance measure. While the LogLoss (equation 1) evaluates the alignment between the predicted probabilities and the actual class (i.e. ORN vs. no ORN), the Brier score (equation 2) is a measure of the overall accuracy of the predicted probabilities (*p*_*t*_) with respect to the actual outcomes (*y*_*t*_) for a total of N predictions. Log Loss and Brier score values may range between 0 and 1 and lower values correspond to a better calibration.

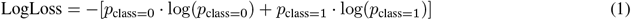

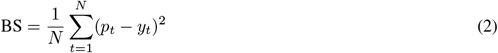

In mis-calibrated models, the alignment between the predicted probabilities and the true class outcome can be adjusted using calibration methods. Platt scaling [29] is a model-agnostic (post-hoc) calibration correction method that can be applied directly to the raw predicted probabilities without involving the model in the process. Platt scaling applies a logistic transformation to the raw predicted probabilities as per equation 3. A logistic regression (LR) model with coefficients A and B is fitted to the raw predicted probabilities f(x) (output from the external validation of the DN40 model on the OUH dataset) to obtain the calibrated probabilities for the positive class *P* (*y* = 1 | *x*). To implement this, the set of raw probabilities was split into training and test subsets following a 5-fold CV approach. The training subset was used to fit the LR model, which was then fitted to the test subset to obtain the calibrated probabilities (Figure 2).

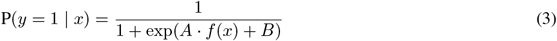

## 3 Results

### 3.1 Dosimetric comparison

Visual representation of the mean DVH for the two datasets (GSTT and OUH) for each group (ORN and control) allowed for an overall assessment of dosimetric differences between groups and datasets. Although the dose distribution varied from patient to patient within each institution (largely depending on the primary treatment site), the mean dose distribution in the GSTT cohort was higher than in the OUH cohort. The GSTT control group received similar doses to the mandible to the OUH ORN group (Figure 3).

**Figure 3:**
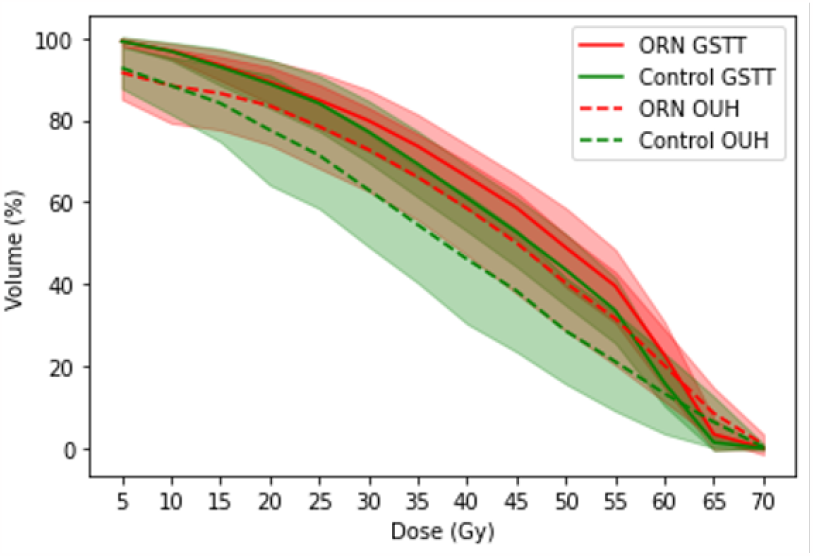
Mean dose-volume data comparison between the OUH and GSTT datasets for the ORN and control groups separately. The shaded areas correspond to the 95% confidence intervals.

### 3.2 Model performance

The DN40 ORN model showed a reasonably good calibration performance at internal validation (Table 2). As shown in the probability frequency histogram in the corresponding plot in Figure 4a, the frequency distribution of the predicted probabilities at internal validation was in line with the true class, i.e. a higher frequency in probabilities under 0.5 was observed in negative cases (class 0). In contrast, positive cases (class 1) obtained a larger proportion of probabilities above 0.5. When testing the model on the independent dataset, i.e. at external validation, calibration performance worsened. As shown in Figure 4b, a clear mismatch between the predicted probabilities and the true classes was observed, with predicted probabilities mostly within the lower range (< 0.5) both for true ORN and control cases. When Platt scaling was applied to the predicted probabilities, a slight shift of the probability range was observed towards values > 0.6 (Figure 4c). However, this did not result in a significant improvement in the calibration metrics. Based on the dose distribution to their mandible, the model’s ability to discriminate between the ORN and no ORN subjects was worse on the independent dataset (see Figure 5 in supplementary material). Platt scaling aims to adjust the probabilities to improve their calibration while preserving the rank order of the predictions (as measured by the ROC AUC). Thus, although Platt scaling improved the balance between recall and specificity at external validation (Table 2), due to the shift in the predicted probabilities range, the overall discrimination performance did not change. The discriminative ability of the RF model trained on DVH metrics (AUROC 0.669) was higher than the DN40 model trained on radiation dose distribution maps (AUROC 0.631). However, this difference was not statistically significant (DeLong p-value 0.667).

**Table 2:** Model calibration and discrimination performance results at internal and external (pre- and post-Platt scaling) validation of the DN40 ORN prediction model. The class probability threshold was set to 0.5 to determine the AUROC.

|  | GSTT dataset | OUH dataset (pre-Platt scaling) | OUH dataset (post-Platt scaling) |
| --- | --- | --- | --- |
| Brier score | 0.229 | 0.239 | 0.239 |
| Intercept | -0.136 | 0.236 | -0.006 |
| Slope | 0.564 | 0.977 | 0.806 |
| Log loss | 0.664 | 0.671 | 0.672 |
| AUROC (95% CI) | 0.693 (0.616-0.769) | 0.631 (0.509-0.753) | 0.631 (0.509-0.754) |
| Accuracy | 0.674 | 0.634 | 0.622 |
| Recall | 0.63 | 0.85 | 0.71 |
| Specificity | 0.72 | 0.42 | 0.54 |
| Precision | 0.69 | 0.59 | 0.60 |

**Figure 4:**
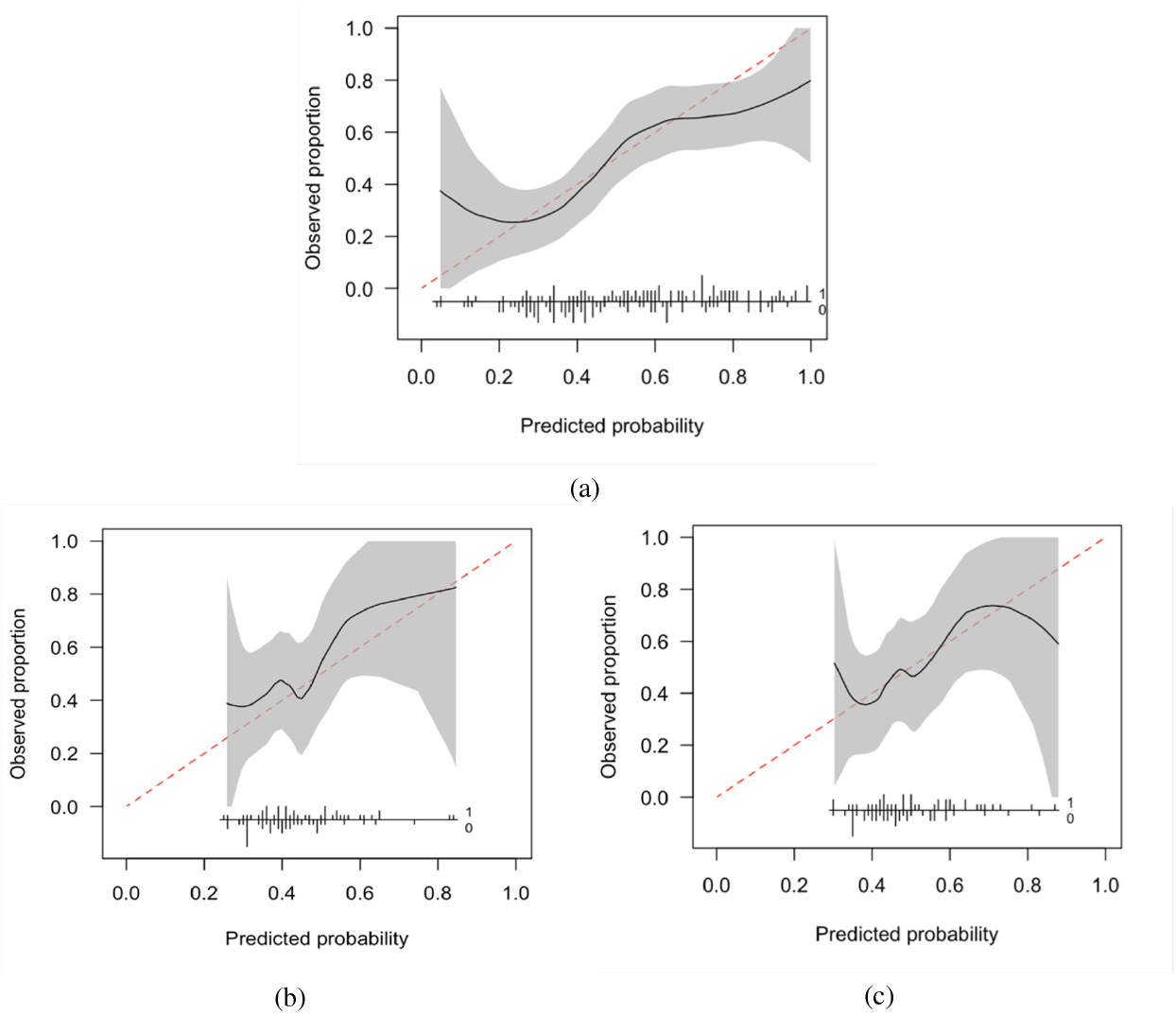
Reliability plots of the DN40 ORN model a) at internal validation b) at external validation before Platt scaling and c) at external validation after Platt scaling. The predicted probabilities were grouped into 10 bins of equal size (i.e. same number of subjects per bin) based on their predicted probability of positive outcome (ORN). The x-axis represents the mean predicted probability of the positive outcome for each bin. The y-axis represents the fraction of true positives (i.e. ORN) within each bin. The shaded area corresponds to the pointwise 95% confidence interval, and the dotted line represents the ideal calibration curve. The relative frequency distribution of the predicted probabilities is plotted at the bottom of each sub-figure with the actual outcome (ORN or no ORN) indicated by the numbers 1 and 0, respectively.

## 4 Discussion

The overall model performance, assessed with the Brier score, was worse at external validation than at internal validation and did not improve after recalibration of the predicted probabilities using Platt scaling. The predicted probabilities on the OUH dataset were largely within the intermediate-to-low range, as the model predicted mostly no ORN cases, which resulted in an increased specificity but worse recall performance. This is most likely because the radiation doses in the mandible were lower overall in the OUH dataset than in the GSTT dataset. Interestingly, however, the tails of the DVH, doses at the higher dose points, were lower for the GSTT DVH, and the average GSTT control doses were similar to the average OUH ORN doses (Figure 3).

Although the discrimination ability of the DN40 model was slightly lower at external validation, this was statistically comparable to that of a DVH-based RF model for the same dataset. This indicates that, in addition to potential model overfitting issues, the small drift in AUROC from internal to external validation was also likely driven by the dosimetric characteristics of the external dataset. The calibration results at external validation, and the lack of improvement after Platt scaling of the predicted probabilities, also indicate that there could be a potential domain shift.

Chemotherapy is known to enhance the sensitivity of tissues to radiation, thus contributing to the development of radiation-induced toxicities. The internal and external cohorts considered in this study differed largely in percentage of patients receiving chemotherapy. Additionally, the difference in chemotherapy rates between the ORN and control group was larger in the internal cohort. These differences between cohorts could have added to the domain shift and thus contributed to the lack of generalisability of the model.

Future work will consider re-training the model on the combined dataset and repeating the external validation exercise on a third independent cohort. The PREDMORN study [22] will provide such an opportunity.

The introduction of regularisation techniques like batch normalisation in the architecture of DL classification neural networks has resulted in deeper and faster networks with improved classification accuracy. However, it has been shown that such models (e.g., DenseNet) are often miscalibrated [30]. Future work will continue to optimise the use of DL in dose map based NTCP models regarding classification accuracy and generalisability by considering alternative neural networks with less regularisation.

We tried to align the predicted probabilities to the true class probabilities with the Platt scaling calibration method. However, in our study, it had only a marginal effect. This calibration method was chosen because it works directly with the raw predicted probabilities and does not require including the model in the calibration process. Isotonic calibration is an alternative model-agnostic method, but it is not as suitable for small datasets [30, 31].

This study investigated the technicalities of translating DL-based toxicity prediction models to external datasets. Thus, a technically convenient class-balanced approach was used to develop and test the DN40 model, and both the internal and external datasets did not represent the real-world ORN prevalence. As a result, the predicted probabilities should not be considered as actual ORN risk predictions.

Finally, in this study, we have only included dose information in the task of predicting mandibular ORN. However, as discussed earlier, other risk factors contribute to this condition. Combining image data, (i.e. radiation dose maps) with tabular clinical data in DL-based NTCP models is not as trivial as in traditional DVH-based NTCP approaches. Different multi-modality data fusion strategies have been suggested, but further work is required to assess the suitability of these in the NTCP modelling domain [32, 33].

## 5 Conclusion

The inclusion of spatial dose information in NTCP models has been the focus of recent research studies, with DL methods proposed as one of the tools to achieve this. External validation of a prediction model is crucial for translating from the research to the clinical setting. It also provides insight into the model’s limitations and potential directions for further development. This is particularly relevant when using DL methods, which are less interpretable than more traditional statistical approaches. Using spatial dose distributions rather than DVH data, NTCP models could provide insight into spatial ORN risk, assisting in dental and oncological management decisions. Our results have highlighted the risk of model overfitting and the importance of domain shift in dose map-based DL models for ORN prediction. Further work is required to develop and test accurate and generalisable spatial ORN models before such individual risk predictions can be used clinically.

## Data Availability

All data produced in the present study are available upon reasonable request to the authors

## Confict of Interest

None.

## Declarations

The GSTT research database (Guy’s Cancer Cohort) used in this study received ethics approval from the North West - Haydock Research Ethics Committee of the NHS Health Research Authority (REC reference 18/NW/0297, IRAS project ID: 231443).

## 6 Acknowledgement

We gratefully acknowledge the support of NVIDIA Corporation with the donation of the Titan Xp GPU used for this research. This work was supported by the Radiation Research Unit at the Cancer Research UK City of London Centre Award [C7893/A28990] and by the Guy’s Cancer Charity via a donation from the Wilson-Olegario foundation and other donations.

## Notes

### Competing Interest Statement

The authors have declared no competing interest.

### Author Declarations

The GSTT research database Guy's Cancer Cohort) used in this study received ethics approval from the North West - Haydock Research Ethics Committee of the NHS Health Research Authority (REC reference 18/NW/0297, IRAS project ID: 231443).

